# A grading system of dynamic fibrinolysis resistance in sepsis associates with ICU outcomes

**DOI:** 10.64898/2026.03.25.26349336

**Authors:** Lucy A. Coupland, Steven A. Frost, Jessica Lin, Nguyen Pham, Eurwin Suryana, Matthew Self, Joanne Chia, Tammy Lam, Zikou Liu, Robert Jaich, Philip J. Crispin, David J. Rabbolini, Ruby H.P. Law, Charithani B. Keragala, Robert L. Medcalf, Anders Aneman

**Author notes:** **Corresponding authors** Prof Anders Aneman: Liverpool Hospital Intensive Care Unit Elizabeth St, Liverpool, NSW, Australia, 2170; Dr Lucy Coupland: Liverpool Hospital Intensive Care Unit Elizabeth St, Liverpool, NSW, Australia, 2170. **Authors Contributions:** LC was involved in study conceptualisation, VET, data collection, analysis and presentation, and manuscript writing; SF was involved in study conceptualisation, data analysis, modelling and presentation, and manuscript writing; JL, NP, ES, MS and RJ were involved in study conceptualisation and data collection; JC, TL and ZL were involved in conceptualising, undertaking and analysing laboratory-based analyses; PC, DR, and RL were involved in study conceptualisation, provided intellectual input and were involved in manuscript editing; CK and RM were involved in study conceptualisation, provided intellectual input, managed and supervised laboratory-based activities and manuscript editing; AA was involved in study conceptualisation, VET, data analysis, modelling and presentation, and manuscript writing. All authors read and approved the final manuscript. **Data Sharing Statement:** Data interaction app links provided in manuscript For original data, please contact corresponding authors.

## Abstract

**Rationale:** Fibrinolysis resistance in sepsis associates with thrombotic burden, multi-organ failure and death. The degrees and dynamics of resistance that associate with mortality in acute sepsis are unknown, and a simple tool to aid clinician interpretation of fibrinolysis measurements is lacking.

**Objectives:** To establish a point of care grading tool of fibrinolysis resistance that aligns with scoring systems for disease acuity, is substantiated by plasma fibrinolysis markers and enables rapid investigation of the fibrinolysis state at the point of care.

**Methods:** Prospective observational study of 116 adult sepsis/septic shock patients with sequential measurements of fibrinolysis resistance during Intensive Care Unit (ICU) admission using tissue plasminogen activator (tPA) enhanced viscoelastic testing (VET). The clot lysis time (TPA-LT) adjusted for fibrin clot amplitude (TPA-LT/FIBA10, sec/mm) underwent cluster analysis and was evaluated against disease severity scores, standard pathology, clinical outcomes and fibrinolysis markers.

**Measurements and Main Results:** Three clusters of progressively increasing fibrinolysis resistance were identified (Grades 1-3). At admission, Grade 3 associated with the highest disease severity, organ failure, haematological and biochemical perturbations, fibrinolysis marker inhibitory profile and mortality (42% versus 24% and 15% in Grade 2 and Grade 1, respectively) with a 3.9-fold [95% CI 1.4-11] increased hazard ratio for death at 28 days compared to Grade 1. Transitions between grades were frequent over 7 days with a reduced Grade associated with decreased risk of death.

**Conclusions:** Grading of fibrinolysis resistance in sepsis enables rapid identification of patients at greatest mortality risk with any dynamic improvement corresponding to favourable clinical outcomes.

## Introduction

Sepsis, defined as organ dysfunction from a dysregulated host response to infection (1), impacts all age groups and causes an estimated 22 million deaths per annum worldwide (2). The responses of the immune, inflammation and coagulation systems in sepsis are highly interlinked with reciprocal amplification mechanisms (3, 4) that lead to widespread generation of microthrombi (5, 6). Reduced fibrinolytic activity in sepsis, referred to as fibrinolysis resistance (7), confers a greater propensity for thrombi to persist and extend (4, 8).

Thrombotic microcirculatory obstruction impairs tissue perfusion contributing to organ dysfunction, which may progress to organ failure and death (6, 9, 10). While a state of impaired fibrinolysis is recognised to play a pivotal role in the coagulopathy associated with sepsis, the degree and dynamics of fibrinolysis in early sepsis remain largely undetermined. Early and prompt recognition of improving or deteriorating fibrinolytic capacity in the initial phase of sepsis is important, since therapeutic interventions are likely to be most effective at this stage before irreversible organ damage has developed.

The contemporary state of fibrinolysis in individual patients can be measured within minutes at the point-of-care using whole blood viscoelastic testing (VET) (11–13). The inclusion of tissue plasminogen activator (tPA) to enhance fibrinolysis within the tissue factor-activated VET (tPA-VET) substantially improves the sensitivity to detect fibrinolysis resistance (14–21). In previous studies using tPA-VET approximately half of patients admitted to the intensive care unit (ICU) for sepsis demonstrated fibrinolysis resistance, and the extent correlated with the severity of sepsis and predicted risk of death (16, 18, 22). These previous studies measured fibrinolysis by tPA-VET at a single point in time (16, 18), thus the evolution over time and the dynamic relationship to outcomes remain unclear. Fibrinolysis resistance may exhibit a range of severity rather than being represented by a discrete state, and the degree that associates with poor patient outcomes is unknown. A grading system that enables bedside clinicians to interpret fibrinolysis resistance is an important step to move evidence in this field towards clinical application.

This study aimed to develop a fibrinolysis resistance grading tool in sepsis for bedside clinical application that uses data readily available from tPA-VET. We hypothesised that this grading system aligns with severity of illness scoring systems for organ failure and coagulopathies, as well as patient outcomes, and be substantiated by plasma levels of key factors influencing fibrinolysis.

## Methods

### Study design and setting

This pragmatic, observational, cohort study was undertaken in the ICU of a tertiary, academic hospital in Liverpool, Australia, with the research program approved by the South Western Sydney Local Health District Human Research Ethics Committee (2022/ETH02122). Consent was waived as permitted under the National Statement on Ethical Conduct in Human Research 2023 for low risk research in critically ill patients. Patients were recruited and VET data collected prospectively, with de-identified clinical and pathology data added retrospectively. The study adheres to the STROBE guidelines for observational studies (23) (Online Data Supplement, Table E1).

### Participants and controls

Adult patients (≥18 years) were eligible for the study if admitted with sepsis or septic shock (according to Sepsis-3 criteria (24)) from a suspected or documented infection, and for active ICU management. Patients with a pre-existing chronic coagulation disorder, known pregnancy, admitted with palliative intent or with the first tPA-VET performed after 72 hrs of admission were excluded. Two control groups were included comprising 26 healthy volunteers recruited among the ICU staff and 32 patients sampled immediately prior to coronary bypasss surgey. The latter group was used to represent comorbid, non-septic patients.

### Clinical data and outcome variables

Data was obtained from patient medical records and included patient characteristics, APACHE III (Acute Physiology and Chronic Health Evaluation), SOFA (Sequential Organ Failure Assessment), SIC (Sepsis Induced Coagulopathy) (25), and DIC (disseminated intravascular coagulation) (26) scores, clinical frailty score (27), biochemistry, haematology, coagulation, arterial blood gas results, organ support measures, steroid and therapeutic anticoagulant administration, and duration and outcome of the ICU admission up to 28 days.

### Viscoelastic measurements and key parameters

ClotPro® (Haemoview Diagnostics, Milton, QLD, Australia) VET point-of-care technology was used to assess coagulation and fibrinolysis. Blood was collected into 3.2% buffered sodium citrate tubes with three assays performed within 30 minutes: (i) EX-test - whole blood coagulation activated with tissue factor with a heparin inhibitor included; (ii) FIB-test - same as the EX-test with two platelet inhibitors included to assess fibrin clot formation; and (iii) TPA-test - same as the EX-test with tPA (650 ng/mL) added to permit evaluation of fibrinolysis. The VET parameters reported include: EX-test clotting time, the time taken for the clot formed to reach 2 mm (EX-CT); the FIB- and EX-test clot amplitudes 10 min after commencement of clot formation (FIB or EXA10, mm); the TPA-test clot lysis time, the time taken for 50% of the maximum clot firmness to be lysed (TPA-LT, sec), TPA-test maximum clot firmness/amplitude was measured during the total assay run time (initially 40 min for 7 patients, then extended to 60 min) (TPA MCF, mm), and the TPA-test maximum clot lysis measured during the assay (TPA ML, %).

### TPA-LT/FIBA10 ratio as an optimised measure of fibrinolysis

In line with the basic principles of enzyme activity, a relationship is seen between the amount of substrate present (fibrin) and the time taken for its enzymic degradation following tPA conversion of plasminogen to plasmin. The amplitude of the fibrin clot (FIBA10), therefore, has a major impact on the clot lysis time (TPA-LT). Significant increases in the FIBA10 are seen in sepsis patients reflecting elevations in plasma fibrinogen as part of the inflammatory response. By spiking increasing concentrations of fibrinogen into healthy control blood, we confirmed that the tPA concentration within the TPA-test (650 ng/mL) is not saturated by the fibrin clot amplitudes seen in sepsis (Online Data Supplement Figure E1). Throughout this paper, therefore, the TPA-LT/FIBA10 ratio (sec/mm), obtained by a simple division, is used as a fibrin-adjusted measure of fibrinolysis. An example of the viscoelastograms from which the TPA LT and FIBA10 values were obtained are provided in Online Data Supplement Figure E2.

### Detection of plasma fibrinolysis markers

In a nested substudy, blood collected in 3.2% buffered sodium citrate was centrifuged at 5000g for 20 min at room temperature to obtain platelet-poor plasma (28) and frozen at minus 80°C. ELISA kits for plasminogen, α2 antiplasmin, tPA activity and PAI-1 activity were obtained from Innovative Research (Novi, MI, USA); IHUPLGKT, IHUA2APLKTT, IHUTPAKT, IHUPAI1KT, respectively. Technozym® PAP complex TC12060 ELISA kits were obtained from Technoclone (Vienna, Austria). Plasma samples were diluted and processed following the manufacturer’s instructions.

### Reversibility of fibrinolysis resistance ex vivo

In a subset of patients, the reversibility of fibrinolysis resistance was tested ex vivo in whole blood sampled at the same time as the patient’s fibrinolysis assessment was undertaken and plasma collected. The standard TPA- (650 ng/mL tPA) and EX- (no tPA included) tests were modified through the addition of the following agents to the analytical cup immediately prior to the addition of blood and assay reagents: i) alteplase (tPA; 650 ng/mL; Boehringer-Ingelheim, Ingelheim, Gemany) thus to a total dose of 1.3 μg/mL in the TPA-test; ii) plasminogen (143 μg/mL; Hyphen Biomed, Neuville-sur-Oise, France); and iii) an inhibitory antibody to alpha 2 antiplasmin (α2AP; 5 μM; Prof Ruby Law, Monash University, Australia).

### Statistical methods

No formal sample size or power calculations were performed given the exploratory nature of this study. Patient characteristics and outcomes are reported using descriptive statistics as appropriate for non-normally distributed data. Single missing variables to calculate the DIC score were replaced by multiple imputation. Correlations were analysed by the Spearman test. Cluster analysis was performed using a Gaussian Mixture Model with Box-Cox transformed data. Plasma fibrinolysis markers were analysed by a linear mixed-effects model using Grade as a fixed effect and individual patients set as random effect. The degree of fibrinolysis resistance were compared across tertiles of APACHE III and SOFA scores, and across SIC and DIC scores. The Aalen-Johansen multistate approach for competing risks generated the probabilities of being discharged alive, remaining or dying in ICU up to 28 days. A multi- state Markov model estimated the probabilities of transitioning between fibrinolysis grades and into the absorbing states of death or discharge alive in the first week. A Cox proportional hazards model assessed the independent association between change in TPA-LT/FIBA10 and risk of dying in ICU. Statistical significance was set at a two-sided p-value of <0.05.

Statistical analyses were performed using R 4.3.3 (R Core Team, 2024). (see also Detailed statistical methods in Online Data Supplement)

## Results

A total of 199 patients with sepsis or septic shock were admitted to ICU during the study period between May 2022 and December 2024 with 116 patients meeting all inclusion and no exclusion criteria (Table 1). Three patients were co-infected with SARS-Cov-2 and septic shock was present in 74/116 (64%) patients. The median time from ICU admission to the first tPA-VET was 10 hours (IQR 4-19). A median of 3 (IQR 1-4) sequential tPA-VET were performed per patient separated by 25 (IQR 23-48) hours.

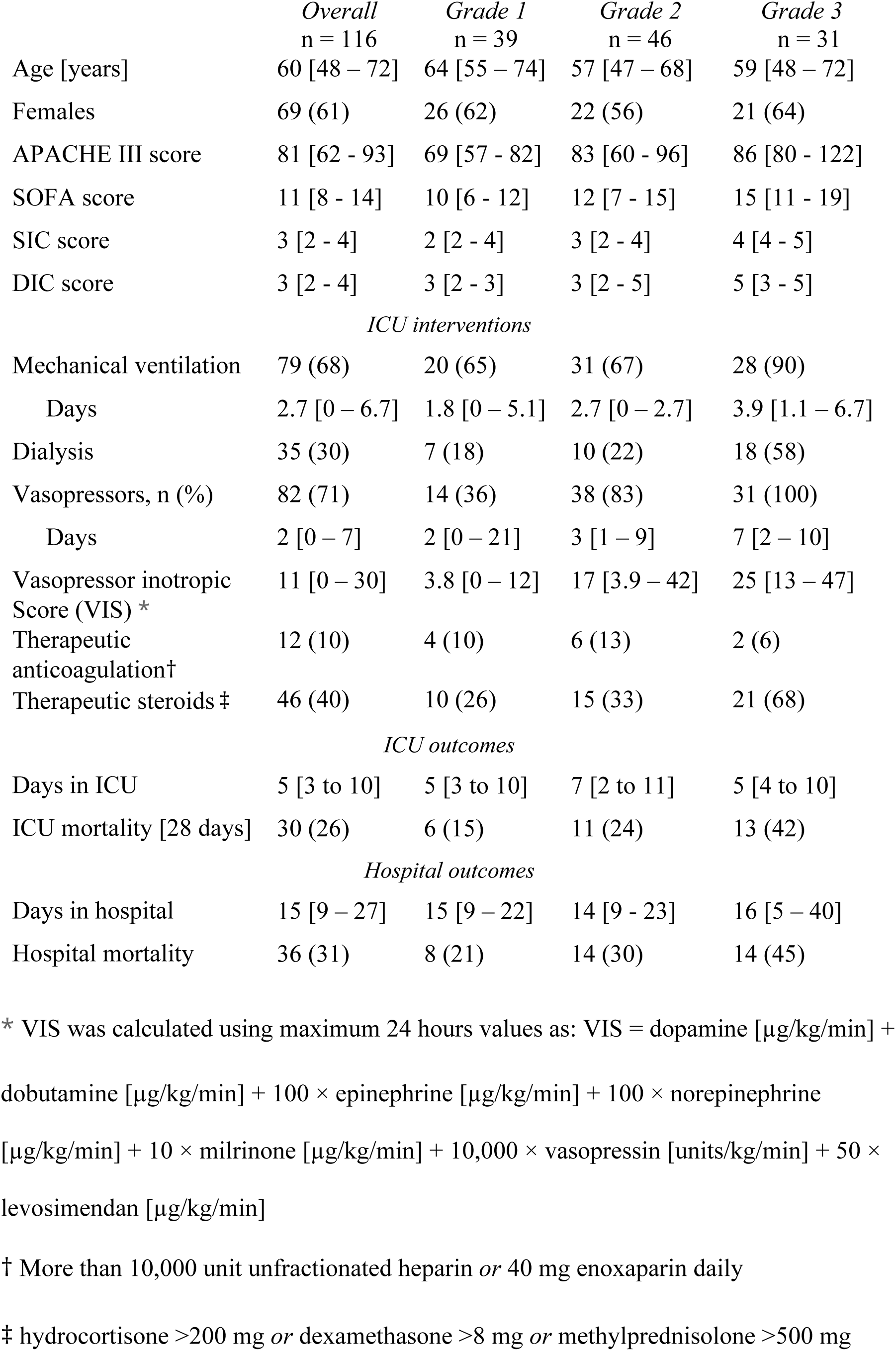

|  | *Overall*<br>n = 116 | *Grade 1*<br>n = 39 | *Grade 2*<br>n = 46 | *Grade 3*<br>n = 31 |
| --- | --- | --- | --- | --- |
| Age [years] | 60 [48 – 72] | 64 [55 – 74] | 57 [47 – 68] | 59 [48 – 72] |
| Females | 69 (61) | 26 (62) | 22 (56) | 21 (64) |
| APACHE III score | 81 [62 - 93] | 69 [57 - 82] | 83 [60 - 96] | 86 [80 - 122] |
| SOFA score | 11 [8 - 14] | 10 [6 - 12] | 12 [7 - 15] | 15 [11 - 19] |
| SIC score | 3 [2 - 4] | 2 [2 - 4] | 3 [2 - 4] | 4 [4 - 5] |
| DIC score | 3 [2 - 4] | 3 [2 - 3] | 3 [2 - 5] | 5 [3 - 5] |
| *ICU interventions* |  |  |  |  |
| Mechanical ventilation | 79 (68) | 20 (65) | 31 (67) | 28 (90) |
| Days | 2.7 [0 – 6.7] | 1.8 [0 – 5.1] | 2.7 [0 – 2.7] | 3.9 [1.1 – 6.7] |
| Dialysis | 35 (30) | 7 (18) | 10 (22) | 18 (58) |
| Vasopressors, n (%) | 82 (71) | 14 (36) | 38 (83) | 31 (100) |
| Days | 2 [0 – 7] | 2 [0 – 21] | 3 [1 – 9] | 7 [2 – 10] |
| Vasopressor inotropic<br>Score (VIS) * | 11 [0 – 30] | 3.8 [0 – 12] | 17 [3.9 – 42] | 25 [13 – 47] |
| Therapeutic<br>anticoagulation† | 12 (10) | 4 (10) | 6 (13) | 2 (6) |
| Therapeutic steroids ‡ | 46 (40) | 10 (26) | 15 (33) | 21 (68) |
| *ICU outcomes* |  |  |  |  |
| Days in ICU | 5 [3 to 10] | 5 [3 to 10] | 7 [2 to 11] | 5 [4 to 10] |
| ICU mortality [28 days] | 30 (26) | 6 (15) | 11 (24) | 13 (42) |
| *Hospital outcomes* |  |  |  |  |
| Days in hospital | 15 [9 – 27] | 15 [9 – 22] | 14 [9 - 23] | 16 [5 – 40] |
| Hospital mortality | 36 (31) | 8 (21) | 14 (30) | 14 (45) |

\* VIS was calculated using maximum 24 hours values as: VIS = dopamine [µg/kg/min] +
dobutamine [µg/kg/min] + 100 × epinephrine [µg/kg/min] + 100 × norepinephrine
[µg/kg/min] + 10 × milrinone [µg/kg/min] + 10,000 × vasopressin [units/kg/min] + 50 ×
levosimendan [µg/kg/min]

### Fibrinolysis Resistance in Controls and Sepsis

The clot lysis time (TPA-LT) and the fibrin clot amplitude (FIBA10) were correlated in healthy controls (r=0.84) including in assays when blood was supplemented with fibrinogen to achieve a greater FIBA10 value than observed in the sepsis patients (Online Data Supplement Figure E1). This correlation was also seen in pre-operative non-septic cardiac patients (r=0.71) (Figure 1A).

**Figure 1.**
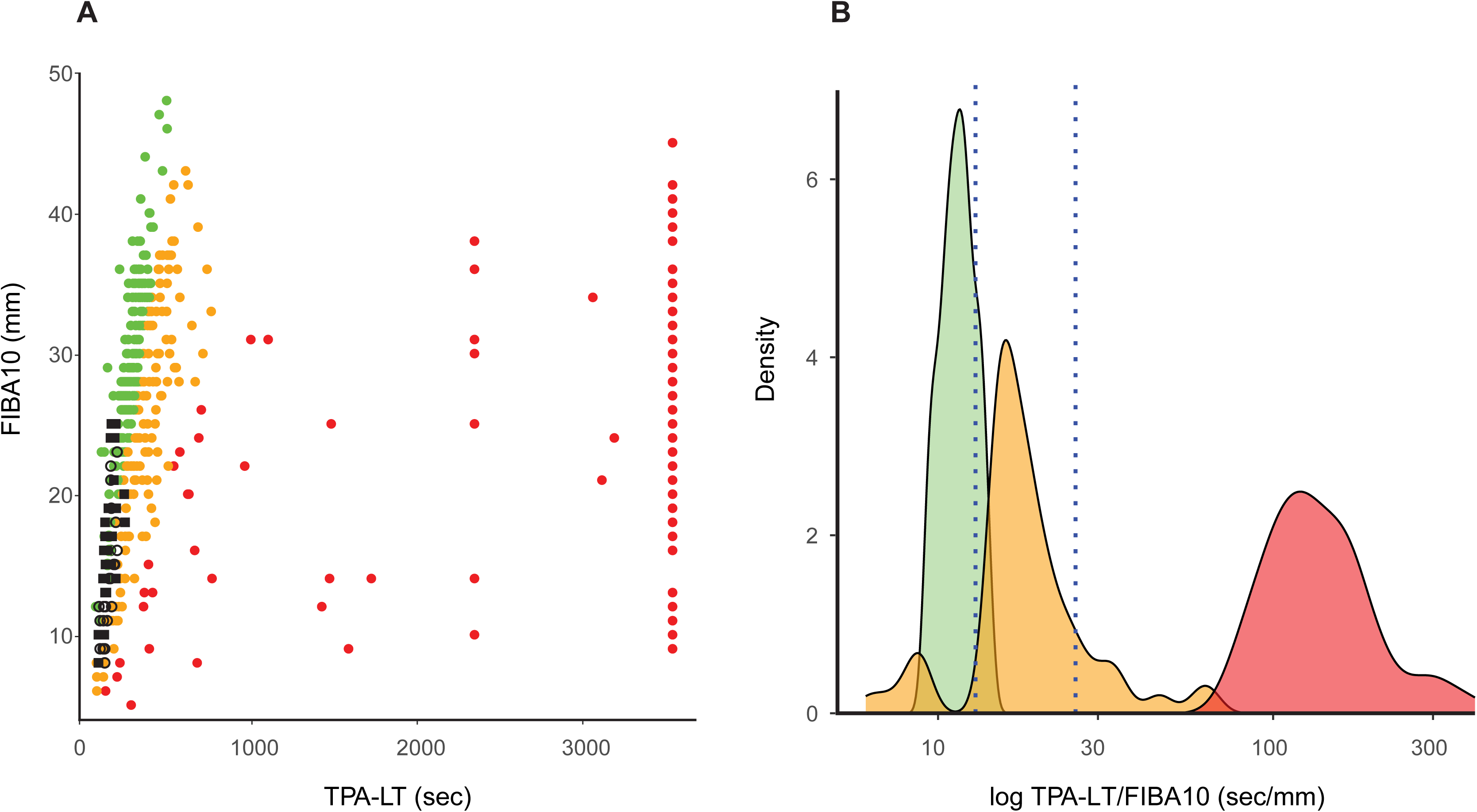
Correlation between clot lysis time (TPA-LT, sec) and fibrin clot formation (FIBA10, mm) (Panel A). Data from healthy subjects (n=26, open black circles), pre-operative cardiac patients (n=32, black solid squares) and sepsis patients (n=116, solid coloured circles). The data driven cluster analysis of all TPA-LT/FIBA10 results identified 3 separate clusters (Panel B) with the numerical thresholds at 13 and 26 sec/mm that were used to define the Grades 1-3 of fibrinolysis resistance. Sepsis patients are coloured according to the TPA-LT/FIBA10 ratio as green (TPA-LT/FIBA10 <13 sec/mm), yellow (TPA-LT/FIBA10 ≥13 and <26 sec/mm) or red (TPA-LT/FIBA10 ≥26 sec/mm).

### Cluster analysis of fibrinolysis resistance severity in sepsis

Three clusters of TPA-LT/FIBA10 ratios were identified in the sepsis cohort, and are referred to as Grades 1-3 with an increasing grade denoting increased fibrinolysis resistance (Figure 1B). Cases with hyperfibrinolysis were rarely seen and were thus not detected in the analysis as an individual cluster. The TPA-LT/FIBA10 <13 sec/mm in Grade 1 included all controls and septic patients with a preserved linear correlation between TPA-LT and FIBA10 (Figure 1A). Grade 2 (TPA-LT/FIBA10 ≥13 to <26 sec/mm) and Grade 3 (TPA-LT/FIBA10 ≥26 sec/mm) reflected increasing fibrinolysis resistance with a gradual loss of the TPA-LT correlation to FIBA10 (Figure 1A; Online Data Supplement Figure E3 and link).

### Fibrinolysis resistance grades, disease severity and ICU support requirements

Patients in Grade 3 fibrinolysis resistance had the highest APACHE III, SOFA, SIC and DIC scores consistent with the most abnormal pathology results, the greatest proportion receiving mechanical ventilation, dialysis and steroids, as well as requiring vasopressor/inotropic agents for a longer duration (Table 1 and Online Data Supplement Table E2). When alternatively analysed by disease severity, tertiles of APACHE III and SOFA scores demonstrated significant association with fibrinolysis resistance severity as measured by the TPA-LT/FIBA10, with a similar association observed with increasing SIC and DIC scores (Online Data Supplement Figure E4).

### Fibrinolysis resistance grades on ICU admission and 28 day outcomes

Patients with Grade 3 fibrinolysis resistance on admission had an increased ICU and in-hospital mortality (Table 1). The hazard ratio for dying in ICU was not significantly increased in Grade 2 fibrinolysis resistance compared to Grade 1, whereas patients in Grade 3 fibrinolysis resistance had a 3.92 fold (95% CI 1.35-11.4) increased hazard ratio (Table 2).

**Table 2.** Hazard ratios for the risk of death in ICU at 28 days.

|  | HR [95% CI] |
| --- | --- |
| Initial grade of fibrinolysis resistance * |  |
| Grade 1 [<13 sec/mm] | 1.0 [reference] |
| Grade 2 [≥13 to <26 sec/mm] | 1.39 [0.47 – 4.11] |
| Grade 3 [≥26 sec/mm] | 3.92 [1.35 – 11.358] |
| Age [each 5-year increase] | 1.07 [0.94 – 1.21] |
| Sex |  |
| Female | 1.0 [reference] |
| Male | 0.52 [0.17 – 1.59] |
| SOFA [each 2 point increase] * | 1.32 [1.02 – 1.73] |
| APACHE III [each 10 point increase] | 1.15 [1.03 – 1.29] |

HR = Hazard Ratio; CI = Confidence Interval; SOFA = Sequential Organ Failure
Assessment; APACHE III = Acute Physiology and Chronic Health Evaluation.
\* Estimates for both TPA-LT/FIBA10 and SOFA scores are based on repeat measures

The Grade 3 fibrinolysis resistance group had a reduced probability of being discharged alive from ICU, and an increased probability of dying within the ICU up to 28 days (Figure 2; Online Data Supplement Table E3).

**Figure 2.**
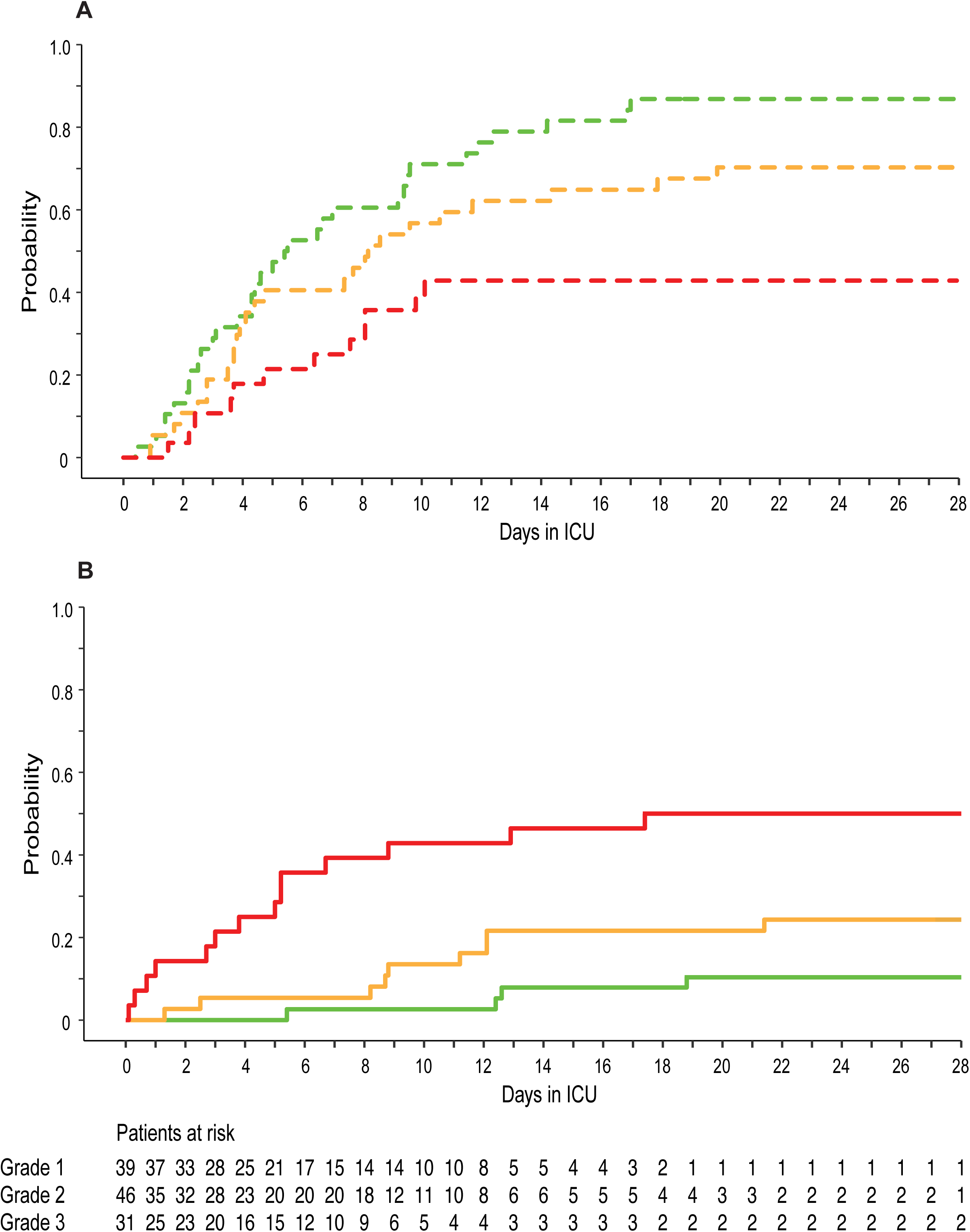
(A) Probabilities of being discharged from ICU alive, or (B) dying in ICU over 28 days. Patients were categorised into Grades 1-3 of fibrinolysis resistance based on the first TPA-LT/FIBA10 measurement in ICU, followed by modelling of 28-day ICU outcomes. Lines are coloured according to the TPA-LT/FIBA10 ratio as green (TPA-LT/FIBA10 <13 sec/mm), orange (TPA-LT/FIBA10 ≥13 and <26 sec/mm) or red (TPA-LT/FIBA10 ≥26 sec/mm). The numbers of patients at risk for either outcome are listed, grouped by initial Grade of fibrinolysis resistance.

### Dynamic transitions between grades and outcomes during the first week in ICU

The predicted risk of ICU death over 7 days was proportionately reduced with improvements in the TPA-LT/FIBA10 (Online Data Supplement Figure E5 and link).

The dynamic transitions between fibrinolysis resistance grades in the first week after admission to ICU are shown in Figure 3. The highest probability of transitioning to death and lowest probability of transitioning to discharge were associated with Grade 3 fibrinolysis resistance at all times.

**Figure 3.**
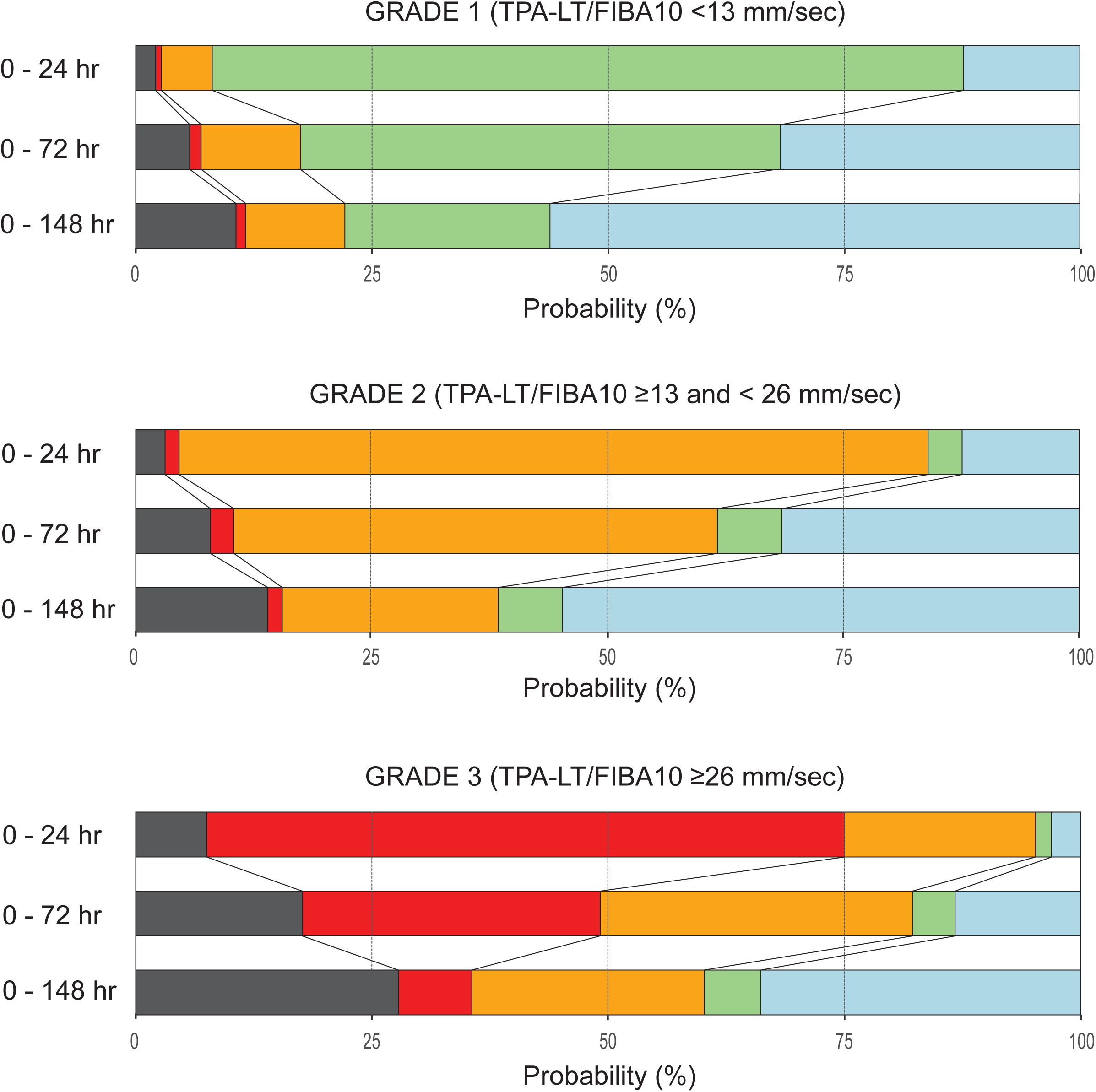
Fibrinolysis resistance Grade transitions and outcome probabilities from start of ICU admission to the end of days 1, 3 and 7. The lengths of the segments in each horizontal bar represent the relative proportion of patients in each state at the end of the different time frames. The three sets of bars represent patients admitted with initial Grade 1 (top panel), Grade 2 (middle panel) and Grade 3 (bottom panel) fibrinolysis resistance. Possible outcomes include remaining in ICU in Grade 1 (green; TPA-LT/FIBA10 <13 sec/mm), Grade 2 (orange; TPA-LT/FIBA10 ≥13) or Grade 3 (red; TPA-LT/FIBA10 ≥26 sec/mm) fibrinolysis resistance or entering the absorbing states of discharged alive (blue) or dying in ICU (black).

### Fibrinolysis Resistance Grades and Plasma Fibrinolysis Proteins

Plasma levels of fibrinolysis proteins were analysed in 162 samples collected sequentially from 40 patients, the majority of whom had transitioned between fibrinolysis resistance grades (8 cases transitioned through 3 grades, 15 transitioned through 2 grades, 9 cases stayed in Grade 1, and 8 cases stayed in Grade 3). Grade 3 fibrinolysis resistance was characterised by higher PAI-1 activity levels, lower plasminogen levels and increased PAI-1/tPA and α2AP/plasminogen ratios. Grade 2 fibrinolysis resistance demonstrated an intermediate trend, while significantly different from Grade 3 (Figure 4A-F, Online Data Supplement Table E4).

**Figure 4.**
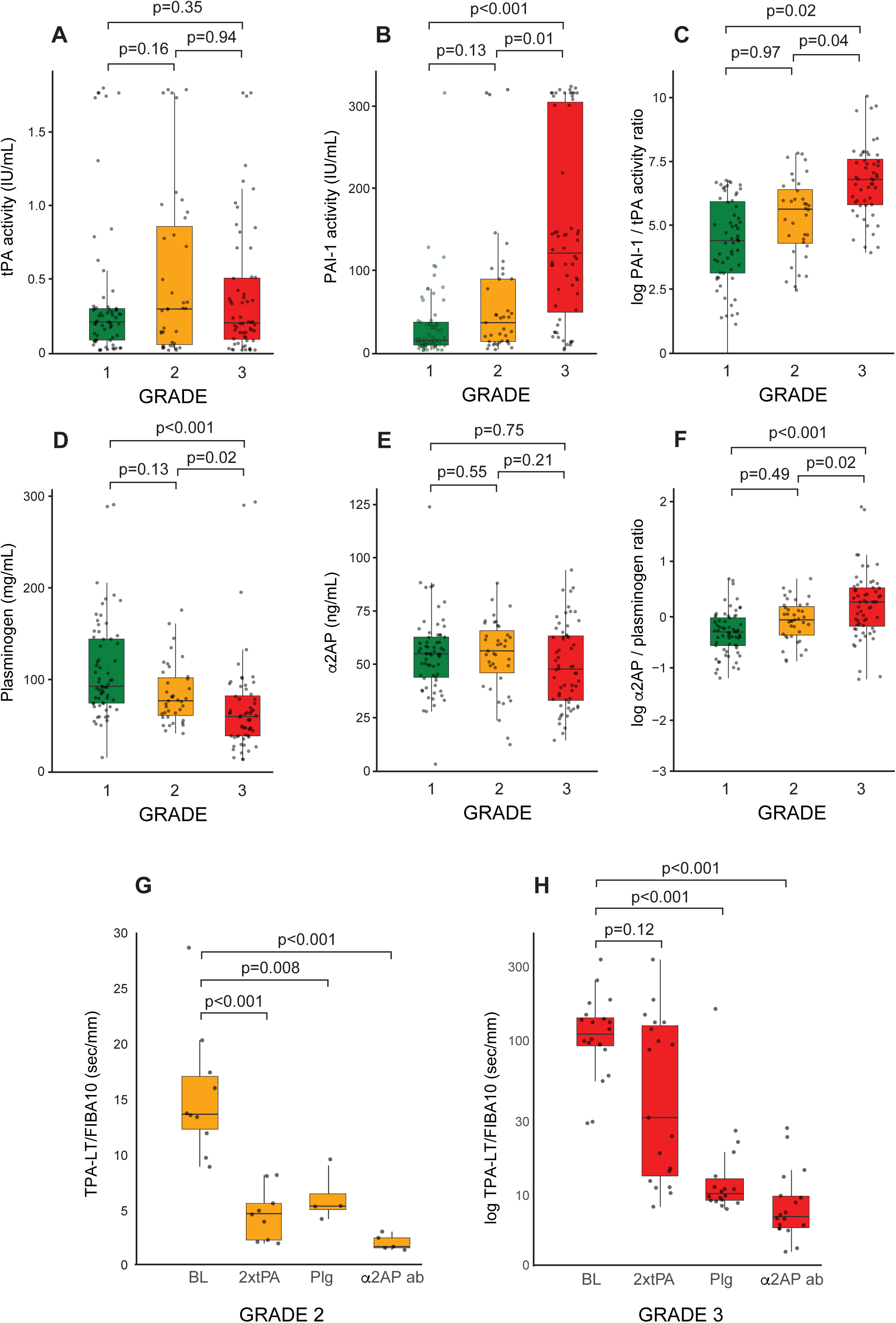
(A-F) Plasma fibrinolysis markers in 40 patients split by fibrinolysis resistance Grades 1-3 (green TPA-LT/FIBA10 <13 sec/mm, Grade 1; yellow TPA-LT/FIBA10 ≥13 and <26 sec/mm, Grade 2; red TPA-LT/FIBA10 ≥26 sec/mm, Grade 3). Note that the y-axes in panels C and F show the 10 log of the ratios. (G-H) The TPA-LT/FIBA10 ratios at baseline and changes following ex vivo supplementation with proteins as indicated. tPA = tissue plasminogen activator; PAI-1 = plasminogen activator inhibitor type 1; α2AP = alpha 2 antiplasmin; BL = baseline; Plg = plasminogen; α2AP ab = inhibitory antibody to alpha 2 antiplasmin.

In the ex vivo reversibility sub-study, in 100% of Grade 2 fibrinolysis resistance cases, a reduction to Grade 1 resistance was achieved by doubling the concentration of tPA in the test, or by adding an α2AP inhibitory antibody to the single dose of tPA (Figure 4G, Online Data Supplement Table E5). In contrast, in Grade 3 resistance samples, doubling the tPA concentration achieved a reduction to Grade 1 in just 26% of cases, to Grade 2 in 21% of cases, while 53% of cases remained in Grade 3 resistance. The addition of plasminogen to

Grade 3 resistance samples reduced 77% of cases to Grade 1 and 17% to Grade 2, while 6% (1 case) remained in Grade 3 resistance. In contrast the α2AP inhibitory antibody reduced 89% of Grade 3 cases to Grade 1 and 11% of cases to Grade 2 fibrinolysis resistance (Figure 4H, Online Data Supplement Table E5). The addition of plasminogen or the α2AP inhibitory antibody was ineffective when added to the EX-test (equivalent test but without tPA) in both Grade 2 and Grade 3 fibrinolysis resistance cases (data not shown).

## Discussion

The principal findings of this study are that (i) data-driven cluster analysis of 429 fibrinolysis measurements in 116 patients admitted to ICU revealed 3 grades of fibrinolysis resistance severity, (ii) the fibrinolysis grade on admission to ICU aligned with the severity of organ failure and coagulopathy, and distinguished the probabilities of being discharged alive or dying in ICU up to 28 days, (iii) transitions from a higher to lower fibrinolysis resistance grade within the first 7 days in ICU associated with improved outcomes, and (iv) the fibrinolysis grades were substantiated by plasma levels of proteins associated with fibrinolysis regulation. A summary of the distinguishing features between the fibrinolysis resistance grades is provided in Table 3.

**Table 3:**
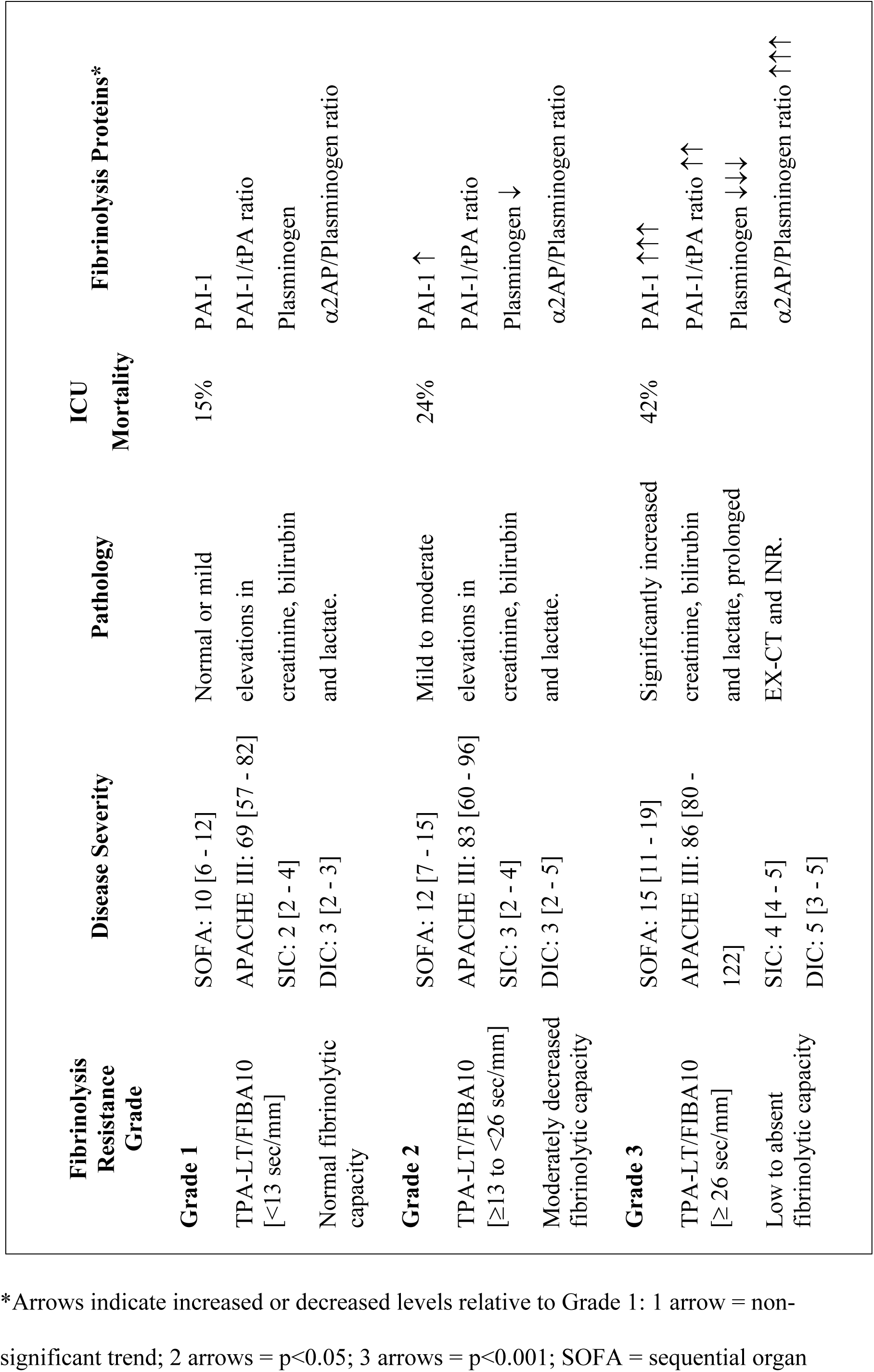
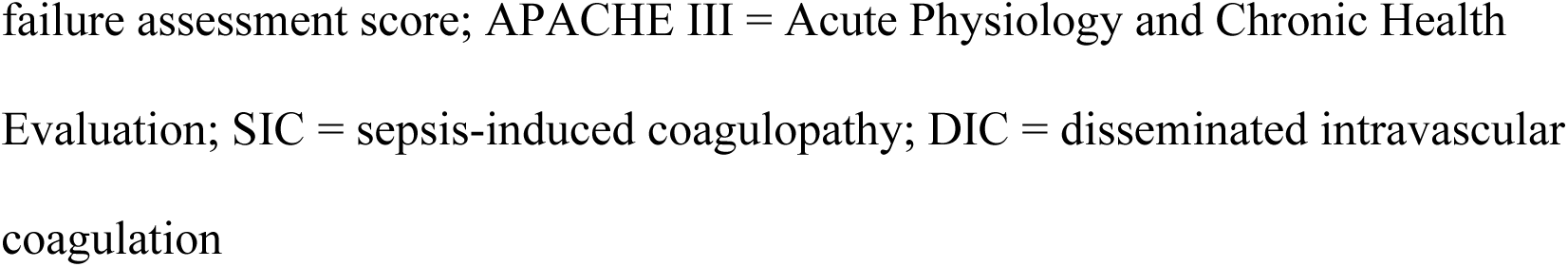
Summary of Grading System for Fibrinolysis Resistance in Sepsis.

| **Fibrinolysis Resistance Grade** | **Disease Severity** | **Pathology** | **ICU Mortality** | **Fibrinolysis Proteins\*** |
| --- | --- | --- | --- | --- |
| **Grade 1** |  |  |  |  |
| TPA-LT/FIBA10 [$<13$ sec/mm] | SOFA: 10 [6 - 12]<br>APACHE III: 69 [57 - 82]<br>SIC: 2 [2 - 4]<br>DIC: 3 [2 - 3] | Normal or mild elevations in creatinine, bilirubin and lactate. | 15% | PAI-1<br>PAI-1/tPA ratio<br>Plasminogen<br>$\alpha 2$AP/Plasminogen ratio |
| Normal fibrinolytic capacity |  |  |  |  |
| **Grade 2** |  |  |  |  |
| TPA-LT/FIBA10 [$\geq 13$ to $<26$ sec/mm] | SOFA: 12 [7 - 15]<br>APACHE III: 83 [60 - 96]<br>SIC: 3 [2 - 4]<br>DIC: 3 [2 - 5] | Mild to moderate elevations in creatinine, bilirubin and lactate. | 24% | PAI-1 ↑<br>PAI-1/tPA ratio<br>Plasminogen ↓<br>$\alpha 2$AP/Plasminogen ratio |
| Moderately decreased fibrinolytic capacity |  |  |  |  |
| **Grade 3** |  |  |  |  |
| TPA-LT/FIBA10 [$\geq 26$ sec/mm] | SOFA: 15 [11 - 19]<br>APACHE III: 86 [80 - 122]<br>SIC: 4 [4 - 5]<br>DIC: 5 [3 - 5] | Significantly increased creatinine, bilirubin and lactate, prolonged EX-CT and INR. | 42% | PAI-1 $\uparrow\uparrow\uparrow$<br>PAI-1/tPA ratio $\uparrow\uparrow$<br>Plasminogen $\downarrow\downarrow\downarrow$<br>$\alpha 2$AP/Plasminogen ratio $\uparrow\uparrow\uparrow$ |
| Low to absent fibrinolytic capacity |  |  |  |  |

\*Arrows indicate increased or decreased levels relative to Grade 1: 1 arrow = non-significant trend; 2 arrows = $p < 0.05$ ; 3 arrows = $p < 0.001$ ; SOFA = sequential organ
failure assessment score; APACHE III = Acute Physiology and Chronic Health Evaluation; SIC = sepsis-induced coagulopathy; DIC = disseminated intravascular coagulation

The tPA-VET used in this study, containing 650 mg/mL tPA, is the most commonly used fibrinolysis enhanced assay reported in the context of sepsis (21). Lower tPA concentrations result in a narrow range of clot lysis within the assay maximum runtime, thus do not distinguish between fibrinolysis resistance grades as was achieved with the higher tPA concentration used in this study (16, 18, 22). By adjusting the clot lysis time for the variation in fibrin clot amplitude, the remaining factors influencing fibrinolysis rate are the concentrations of fibrinolysis activators and their inhibitors. The strong TPA-LT to FIBA10 correlation seen in Grade 1 fibrinolysis resistance reflects the dynamic balance of fibrinolysis seen in healthy controls that contributes to the maintenance of haemostasis. Grade 1 resistance was also observed in non-septic patients with cardiovascular, respiratory and metabolic morbidities, these conditions being prevalent in patients with sepsis. The finding of Grade 1 resistance in septic patients, illustrates that fibrinolysis balance can be maintained in the presence of infection and elevated fibrinogen levels. Importantly, this balance was increasingly disrupted in Grades 2 and 3 resistance which associated with worsening sepsis and septic shock.

The objective data driven cluster analysis demonstrated distinct separation between Grades 2 and 3, correlating with the separation in clinical and outcome data, thus supporting that the grades represent different states with minimal overlap. The finding of poorer outcomes in Grade 3 fibrinolysis resistance agrees with other studies in sepsis patients that demonstrated an association between early impaired fibrinolysis, an increased risk of ICU death, and worsening organ dysfunction (16). A close to a three-fold increased in 30-day mortality was reported in ICU patients with impaired, as compared to normal fibrinolysis (18). Grade 1 and Grade 2 fibrinolysis resistance groups had a lower risk of dying in ICU without a significant difference between the two. Grade 1 may therefore be considered normal fibrinolysis resistance, and Grade 2 a warning of a worsening state, although without the severe clinical implications of Grade 3. The dynamic grades of fibrinolysis resistance presented in this study and their association with different clinical outcomes challenge the prevailing idea that fibrinolysis resistance is a uniform, static feature of sepsis.

A novel and key aspect of this study is the repeated measurements during the ICU admission that allowed the dynamics of fibrinolysis resistance in sepsis to be investigated. These analyses demonstrated that within the first week in ICU, the probabilities of improving through transition from Grade 3 to 2 fibrinolysis resistance were ∼10 fold higher than the probabilities of worsening by moving in the inverse direction. In contrast, patients who had remained or moved into severe Grade 3 fibrinolysis resistance by Day 7 in ICU had the worst ICU outcomes. These findings highlight the potential benefit of monitoring fibrinolysis as an additional real-time indicator that may forecast clinical changes.

Sepsis leads to excessive thrombin generation (29) and deposition of dense fibrin networks which are reported to assist pathogen sequestration (3, 5). At the same time, fibrinolysis is reduced by increased production of the fibrinolysis inhibitors PAI-1 and α2AP (30–32). In contrast the levels of thrombin-activatable fibrinolysis inhibitor (TAFI) are reduced in sepsis (33, 34). Plasminogen levels may become depleted due to an initial hyperfibrinolysis response early in infection, involvement in immune and inflammation modulation and tissue repair (35, 36), and compromised hepatic function (37). Constituents of neutrophil extracelluar traps released in sepsis (38) also degrade plasminogen thus reducing plasmin formation (39).

Overall, this amounts to a prothrombotic and fibrinolysis resistant state. The analyses of fibrinolysis markers in this study aligned with these previous reports. Patients with Grade 3 fibrinolysis resistance had the highest fibrinolysis inhibitory profile driven principally by elevated PAI-1 and decreased plasminogen. The combination of standard dose tPA and α2AP inhibition was effective in restoring Grade 1 resistance in cases of both Grade 2 and Grade 3 resistance. These findings suggest that elevated PAI-1 levels are counterbalanced by tPA supplementation, and α2AP inhibition increases plasmin availability, thus effectively compensating for reduced plasminogen levels.

Future studies are needed to validate the described fibrinolysis grading system in larger sepsis cohorts. In addition to aiding prognostication, it is possible that the grading system could identify in which patients and at what time therapeutic interventions targeting fibrinolysis might be beneficial. The proposed grading system is conducive to enrichment strategies in clinical trials targeting the thrombotic coagulopathy occuring in sepsis and may direct interventions to specific clinical phenotypes (40), thus breaking the notable lack of progress in this area (41, 42).

This study has important strengths and limitations. Point-of-care tPA-VET provides real-time results for the critically ill patient and is easily repeated. The simple calculation of the TPA-LT/FIBA10 ratio enables a more insightful evaluation of factors influencing fibrinolysis. The grading system was established using data-driven cluster analysis. The associations of fibrinolysis resistance with patient outcomes and known plasma mediators of the fibrinolytic system support its clinical relevance and scientific rationale. Fibrinogen and D-dimer tests were requested at the discretion of the treating ICU team thus reducing the available DIC score dataset necessitating the use of data imputation. We acknowledge that other indices of fibrinolysis from tPA-VET have been suggested and could generate different results.

In conclusion, clot lysis time measured in septic patients using tPA (650ng/ml)-enhanced VET on admission to ICU detected three distinct fibrinolysis resistance endotypes that differed in the disease severity, day 28 outcomes, and fibrinolysis marker profiles. Transition to a lower fibrinolysis resistance grade predicted improved outcomes in high risk patients, whereas persistent high grade fibrinolysis resistance was associated with the greatest risk for death in ICU. The proposed fibrinolysis resistance grading system is intended to identify high risk patients, assist clinician management, and aid in progressing interventional studies.

## Data Availability

Data interaction app links provided in manuscript
For original data, please contact corresponding authors.

https://fortitude.shinyapps.io/TPA_FIB_CORREL

https://fortitude.shinyapps.io/CHANGE_TPAFIB

## Acknowledgements

Liverpool Hospital ICU ClotPro Champions – Intensive Care nursing staff who were trained on the ClotPro technology during this study and provided 24/7 capability in the latter half: Alexandra Heath, Emma Hill, Marcus Iedani, Karla Lopez, Elisa McClure, Jennifer McClure, Yvette McManus, Carlos Mendoza, Jennene Miller, Mostafa Hasha, Sharyn Parker, Julio Ruiz Davila, Krystle-Lee Strickland, Anne Tran, Bija Unnikrishnan, Harley Whitfield, Sanjay Singh, Katie Mills. Violeta Jardin and Tony Ghent, Haemoview Diagnostics, Australia, for their technical support and scientific advice. Prof John Myburgh for his critical review of the manuscript.

## List of Abbreviations

α2AP: alpha-2 antiplasmin
APACHE: Acute physiology and chronic health evaluation
DIC: Disseminated intravascular coagulation
EXA10: Clot amplitude 10 min after clot commencement on Ex-test
FIBA10: Clot amplitude 10 min after clot commencement on FIB-test
ICU: Intensive Care Unit
INR: International normalised ratio
IQR: Interquartile range
LT: Clot lysis time
PAI-1: Plasminogen activator inhibitor
PAP: Plasmin antiplasmin complex
PT: Prothrombin time
SIC: Sepsis induced coagulopathy
SOFA: Sequential organ failure assessment
TAFI: Thrombin activatable fibrinolysis inhibitor
tPA: Tissue plasminogen activator
TPA-LT: Clot lysis time obtained from TPA-test
TPA-LT/FIBA10: TPA-LT adjusted for fibrin clot amplitude (FIBA10)
tPA-VET: Tissue plasminogen activator enhanced viscoelastic testing
VET: Viscoelastic testing

