## Supplement for "A grading system of dynamic fibrinolysis resistance in sepsis associates with ICU outcomes"

**Table E1.** STROBE statement.

| | Item No | Recommendation | Page No |
|---|---|---|---|
| **Title and abstract** | 1 | (*a*) Indicate the study's design with a commonly used term in the title or the abstract<br>(*b*) Provide in the abstract an informative and balanced summary of what was done and what was found | Title Page, line 1<br>1, lines 72-84 |
| **Introduction** | | | |
| Background/rationale | 2 | Explain the scientific background and rationale for the investigation being reported | 3-4, lines 90-116 |
| Objectives | 3 | State specific objectives, including any prespecified hypotheses | 4, lines 118-122 |
| **Methods** | | | |
| Study design | 4 | Present key elements of study design early in the paper | 4, line 124 |
| Setting | 5 | Describe the setting, locations, and relevant dates, including periods of recruitment, exposure, follow-up, and data collection | 4, lines 124-132<br>8, line 222 |
| Participants | 6 | (*a*) Give the eligibility criteria, and the sources and methods of selection of participants. Describe methods of follow-up<br>(*b*) For matched studies, give matching criteria and number of exposed and unexposed | 4-5, lines 136-142 |
| Variables | 7 | Clearly define all outcomes, exposures, predictors, potential confounders, and effect modifiers. Give diagnostic criteria, if applicable | 5-7, lines 146-200 |
| Data sources/ measurement | 8* | For each variable of interest, give sources of data and details of methods of assessment (measurement). Describe comparability of assessment methods if there is more than one group | 5-7, lines 146-200<br>Online Supplemental Data |
| Bias | 9 | Describe any efforts to address potential sources of bias | 4, lines 136-143<br>7, lines 203-218<br>Online Supplemental Statistics |
| Study size | 10 | Explain how the study size was arrived at | 7, lines 203-204 |
| Quantitative variables | 11 | Explain how quantitative variables were handled in the analyses. If applicable, describe which groupings were chosen and why | 7, lines 205-217<br>Online Supplemental Statistics |
| Statistical methods | 12 | (*a*) Describe all statistical methods, including those used to control for confounding<br>(*b*) Describe any methods used to examine subgroups and interactions | 7, lines 203-217 |

| | | | |
|---|---|---|---|
| | | (*c*) Explain how missing data were addressed<br>(*d*) If applicable, explain how loss to follow-up was addressed<br>(*e*) Describe any sensitivity analyses | Online Supplemental Statistics |
| **Results** | | | |
| Participants | 13 | (a) Report numbers of individuals at each stage of study—eg numbers potentially eligible, examined for eligibility, confirmed eligible, included in the study, completing follow-up, and analysed<br>(b) Give reasons for non-participation at each stage<br>(c) Consider use of a flow diagram | 8, line 221 - 225 |
| Descriptive data | 14 | (a) Give characteristics of study participants (eg demographic, clinical, social) and information on exposures and potential confounders<br>(b) Indicate number of participants with missing data for each variable of interest<br>(c) Summarise follow-up time (eg, average and total amount) | Table 1 Online Data Supplement |
| Outcome data | 15 | Report numbers of outcome events or summary measures over time | 8-11, lines 229 – 293 Online Data Supplement |

**Table E2.** Patient characteristics and biochemistry results grouped by ICU admission fibrinolysis resistance Grade (1–3). Values are counts (percentages) or median [interquartile range]. Normal laboratory ranges are indicated in the first column

| | **Grade 1** *n = 39* | **Grade 2** *n = 46* | **Grade 3** *n = 31* | **All** *n = 116* |
|---|---|---|---|---|
| Diabetes | 14 (36) | 16 (35) | 7 (23) | 37 (32) |
| Chronic cardiovascular disease | 6 (16) | 5 (10) | 3 (10) | 14 (12) |
| Chronic respiratory disease | 4 (10) | 6 (14) | 2 (6) | 12 (10) |
| Chronic renal disease | 0 (0) | 1 (2) | 2 (6) | 3 (3) |
| Neutro/thrombocytopaenia | 4 (10) | 1 (2) | 3 (10) | 8 (7) |
| Frailty | | | | |
| 1–3 | 11 (29) | 11 (24) | 12 (38) | 34 (29) |
| 4–6 | 25 (65) | 33 (71) | 18 (57) | 76 (66) |
| 7–8 | 3 (7) | 2 (4) | 1 (4) | 6 (5) |
| Infection source | | | | |
| Abdo | 5 (13) | 3 (7) | 9 (29) | 17 (15) |
| Uro | 5 (13) | 5 (10) | 3 (10) | 13 (11) |
| Resp | 11 (29) | 15 (33) | 6 (19) | 34 (29) |
| Soft tissue | 5 (13) | 6 (14) | 6 (19) | 17 (15) |
| Other | 13 (33) | 17 (37) | 7 (23) | 37 (32) |
| Hb, g/L (120–150) | 113 [94–122] | 103 [87–115] | 105 [96–130] | 105 [87–119] |
| White Cells, $10^9$/L (4–10) | 11 [7–17] | 11 [7–14] | 13 [8–19] | 10 [3–6] |
| CRP, mg/L (≤5) | 88 [42–153] | 261 [163–325] | 289 [136–337] | 172 [74–287] |
| Urea, mmol/L (3.5–8.0) | 9 [8–15] | 10 [7–18] | 14 [9–20] | 10 [7–18] |
| Creatinine, µmol/L (45–90) | 106 [69–148] | 137 [70–208] | 198 [113–218] | 114 [66–202] |

| | | | | |
|---|---|---|---|---|
| Bilirubin, µmol/L (≤20) | 10 [5–36] | 13 [10–19] | 33 [23–66] | 12 [6–31] |
| Lactate, mmol/L (≤1.9) | 2.0 [1.6–3.9] | 2.0 [1.0–3.8] | 6.0 [4.0–7.0] | 2.9 [1.7–4.7] |
| Albumin, g/L (30–44) | 23 [21– 35] | 21 [17–23] | 20 [18–22] | 22 [19–24] |
| EX CT, sec (38–65) | 70 [63–80] | 74 [64–85] | 92 [79–123] | 76 [65–95] |
| EX A10, mm (47–64) | 58 [51–63] | 64 [60–67] | 61 [53–65] | 62 [53–65] |
| FIB A10, mm (7–23) | 22 [16–27] | 30 [25–35] | 26 [21–31] | 26 [22–31] |
| PT, sec (9–13) | 15 [13–16] | 15 [12–17] | 18 [13– 19] | 15 [12 – 17] |
| INR (0.9–1.2) | 1.3 [1.2–1.7] | 1.3 [1.1 – 1.5] | 1.6 [1.2 – 2.4] | 1.4 [1.2 – 1.7] |
| D–dimer, mg/L (≤0.5) | 5 (2.2–19) *n=18* | 2.4 (1.9 – 7.3) *n=25* | 2.8 (2.2 – 5.8) (*n=18)* | 3.8 (2.1 – 7.4) |
| Platelets, $10^9$/L (150–400) | 167 [68 – 242] | 195 [157 – 305] | 117 [95 – 204] | 188 [106 – 256] |

EX = EX-test on ClotPro®.

FIB = FIB-test on ClotPro®.

**Table E3.** Probabilities (percentages with 95% confidence intervals) for the outcomes remaining alive in ICU, being discharged alive from ICU or dying in ICU up to 28 days after admission.

| | **Grade 1** | **Grade 2** | **Grade 3** |
|---|---|---|---|
| **Remaining alive in ICU** | | | |
| Day 1 | 95 (89-100) | 95 (88-100) | 88 (77-100) |
| Day 3 | 67 (54-83) | 78 (66-92) | 73 (59-90) |
| Day 7 | 33 (22-51) | 58 (44-75) | 39 (26-60) |
| Day 10 | 24 (14-41) | 33 (21-51) | 24 (13-44) |
| Day 14 | 12 (5.2-27) | 15 (7.2-31) | 15 (6.8-24) |
| Day 21 | 2.4 (0.3-17) | 7.5 (2.5-22) | 9.1 (3.1-27) |
| Day 28 | 2.4 (0.3-17) | 2.5 (0.4-17) | 9.1 (3.1-27) |
| **Discharge alive from ICU** | | | |
| Day 1 | 4.8 (1.2-18) | 5.0 (1.3-19) | 0 (0-0) |
| Day 3 | 33 (22-51) | 18 (8.9-34) | 9.1 (3.1-27) |
| Day 7 | 64 (51-81) | 38 (25-56) | 27 (16-48) |
| Day 10 | 74 (62-88) | 55 (42-73) | 39 (26-60) |
| Day 14 | 81 (70-94) | 65 (52-82) | 45 (31-66) |
| Day 21 | 88 (79-98) | 73 (60-88) | 48 (34-69) |
| Day 28 | 88 (79-98) | 75 (63-90) | 48 (34-69) |
| **Dying in ICU** | | | |
| Day 1 | 0 (0-0) | 0 (0-0) | 12 (4.8-30) |
| Day 3 | 0 (0-0) | 5.0 (1.3-19) | 18 (8.8-37) |
| Day 7 | 2.4 (0.3-17) | 5.0 (1.3-19) | 33 (21-54) |
| Day 10 | 2.4 (0.3-17) | 13 (5.5-28) | 36 (23-57) |
| Day 14 | 7.1 (2.4-21) | 20 (11-37) | 39 (26-60) |
| Day 21 | 9.5 (3.7-24) | 20 (11-37) | 42 (29-63) |
| Day 28 | 9.5 (3.7-24) | 23 (13-40) | 42 (29-63) |

**Table E4.** Plasma levels of fibrinolysis proteins measured in 162 samples from 40 patients and split by TPA-LT/FIBA10 ratio Grade 1-3. Values are median [IQR].

| | **Grade 1** | **Grade 2** | **Grade 3** | **All** |
|---|---|---|---|---|
| **tPA activity** | 0.20 [0.08-0.29] | 0.29 [0.05-0.85] | 0.19 [0.08-0.5] | 0.2 [0.07-0.43] |
| **PAI-1 activity** | 13 [7.9-36] | 35 [12-88] | 120 [48-304] | 35 [11-109] |
| **PAI-1/tPA ratio** | 79 [22-363] | 269 [71-581] | 858 [319-1904] | 308 [62-764] |
| **Plasminogen [Plg]** | 93 [74-145] | 77 [61-102] | 60 [39-83] | 79 [57-108] |
| **α2 antiplasmin [α2AP]** | 55 [44-63] | 57 [46-66] | 48 [33-64] | 55 [39-64] |
| **α2AP/Plg ratio** | 0.52 [0.4-0.67] | 0.65 [0.49-0.83] | 0.9 [0.58-1.17] | 0.63 [0.48-0.90] |
| **Plasmin-antiplasmin complexes** | 628 [507-904] | 626 [538-849] | 579 [358-807] | 616 [455-838] |

**Table E5.** Ex vivo reversibility studies in patients with baseline Grade 2 or Grade 3 TPA-LT/FIBA10 ratios. Values are median [IQR].

| **Grade 2 (≥13 TPA-LT/FIBA10 <26)** | | | |
|---|---|---|---|
| **Baseline** | **2 x tPA** | **Plasminogen** | **α2AP inhibitory ab** |
| 19 [16-23] | 9.6 [7.1-12] | 10 [9.4-14] | 6.6 [6.4-7.7] |
| **Grade 3 (TPA-LT/FIBA10 <26)** | | | |
| **Baseline** | **2 x tPA** | **Plasminogen** | **α2AP inhibitory ab** |
| 108 [87-142] | 31 [12-129] | 10 [9.0-14] | 7.1 [6.0-11] |

**Figure E1.** Correlation between lysis time (TPA-LT) and fibrinogen clot amplitude at 10 minutes (FIBA10) with increasing concentration of fibrinogen added to healthy control blood sample. The linear correlation was $r^2$=0.95.

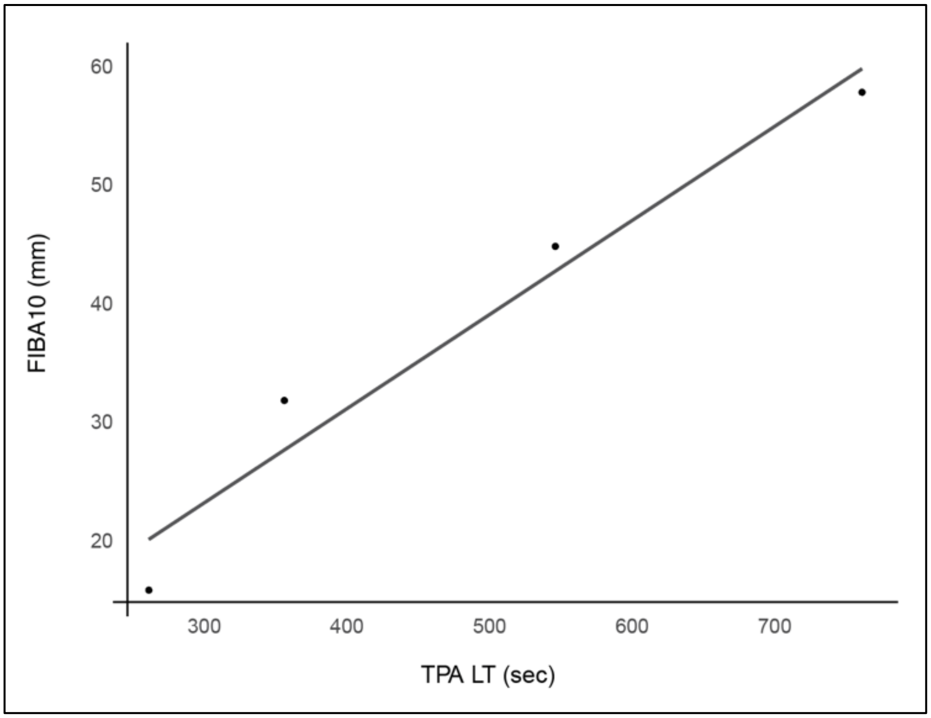

**Figure E2.** Examples of tPA-VET output and calculation of the TPA-LT/FIBA10 ratio.

(A) TPA-LT / FIBA10 = 540 sec / 37 mm = 14.6 sec/mm (Grade 2 fibrinolysis resistance).

(B) When no lysis time is recorded due to a reduction of 50% in the MCF not occurring, a lysis time of 3600 sec should be used, the maximum run time of the assay:-

TPA-LT / FIBA10 = 3600 sec /14 mm = 257 sec/mm (Grade 3 fibrinolysis resistance)

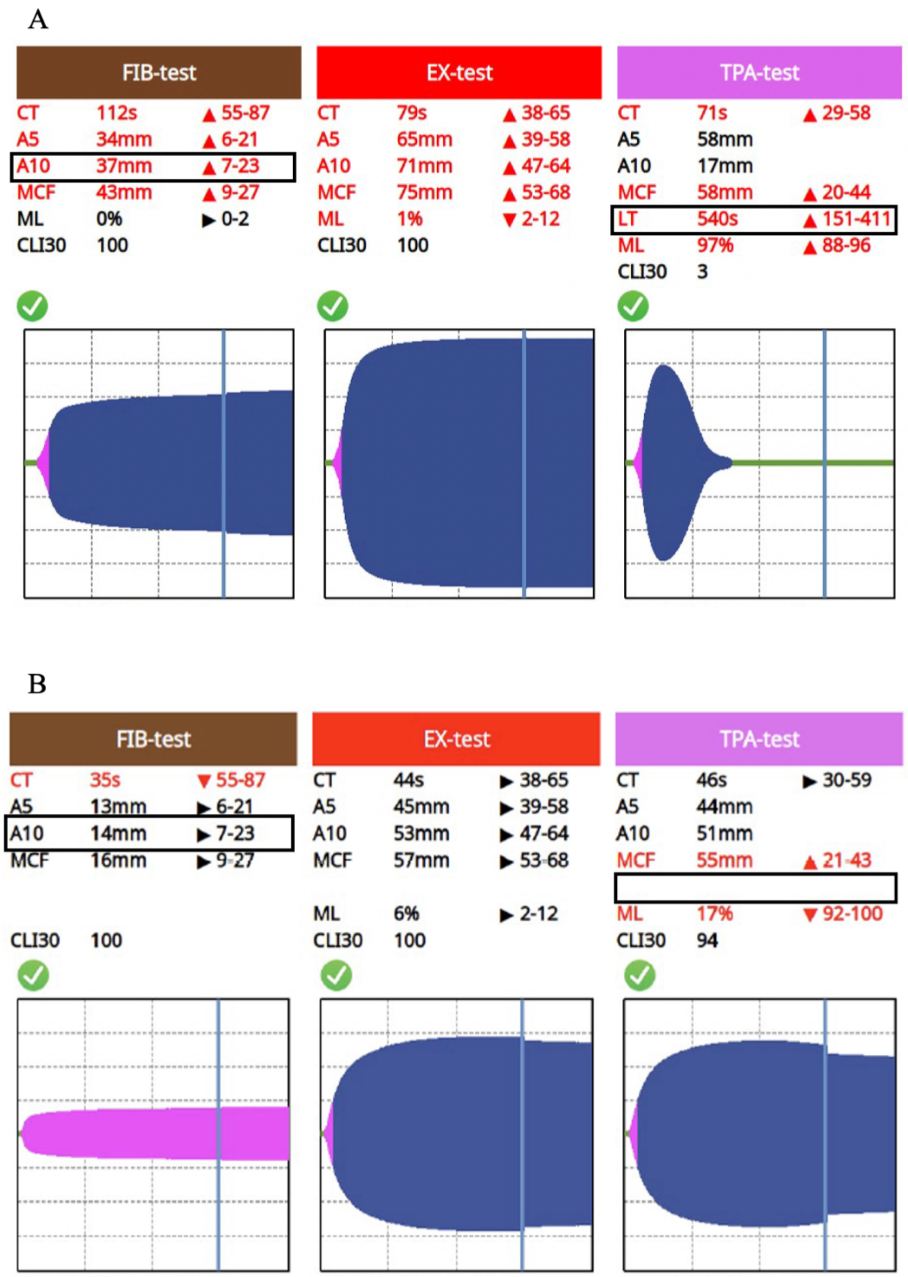

**Figure E3.** The variable correlation between lysis time (TPA-LT) and fibrinogen clot amplitude at 10 minutes (FIBA10) can be explored in the interactive data tool available at: https://fortitude.shinyapps.io/TPA_FIB_CORREL. Data shown are based on the 116 patients, 26 healthy volunteers (open circles) and 32 preoperative cardiac patients (squares).

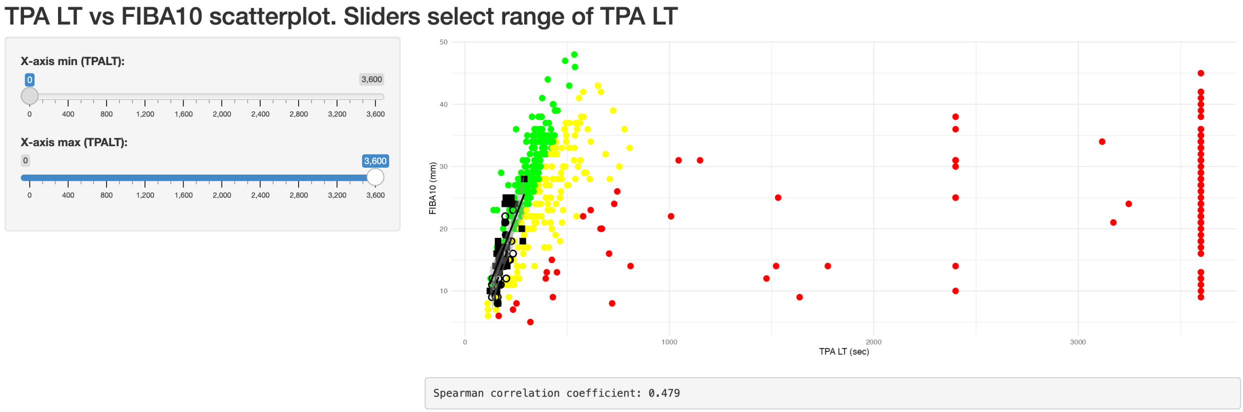


The two sliders on the left select the lower range (top) and upper range (bottom) of TPA-LT to be visualised with a display of the correlation to FIBA10.

**Figure E4.** The TPA-LT/FIBA10 ratio as a measure of fibrinolysis resistance across tertiles of (A) APACHE III, (B) SOFA, (C) SIC and (D) DIC scores.

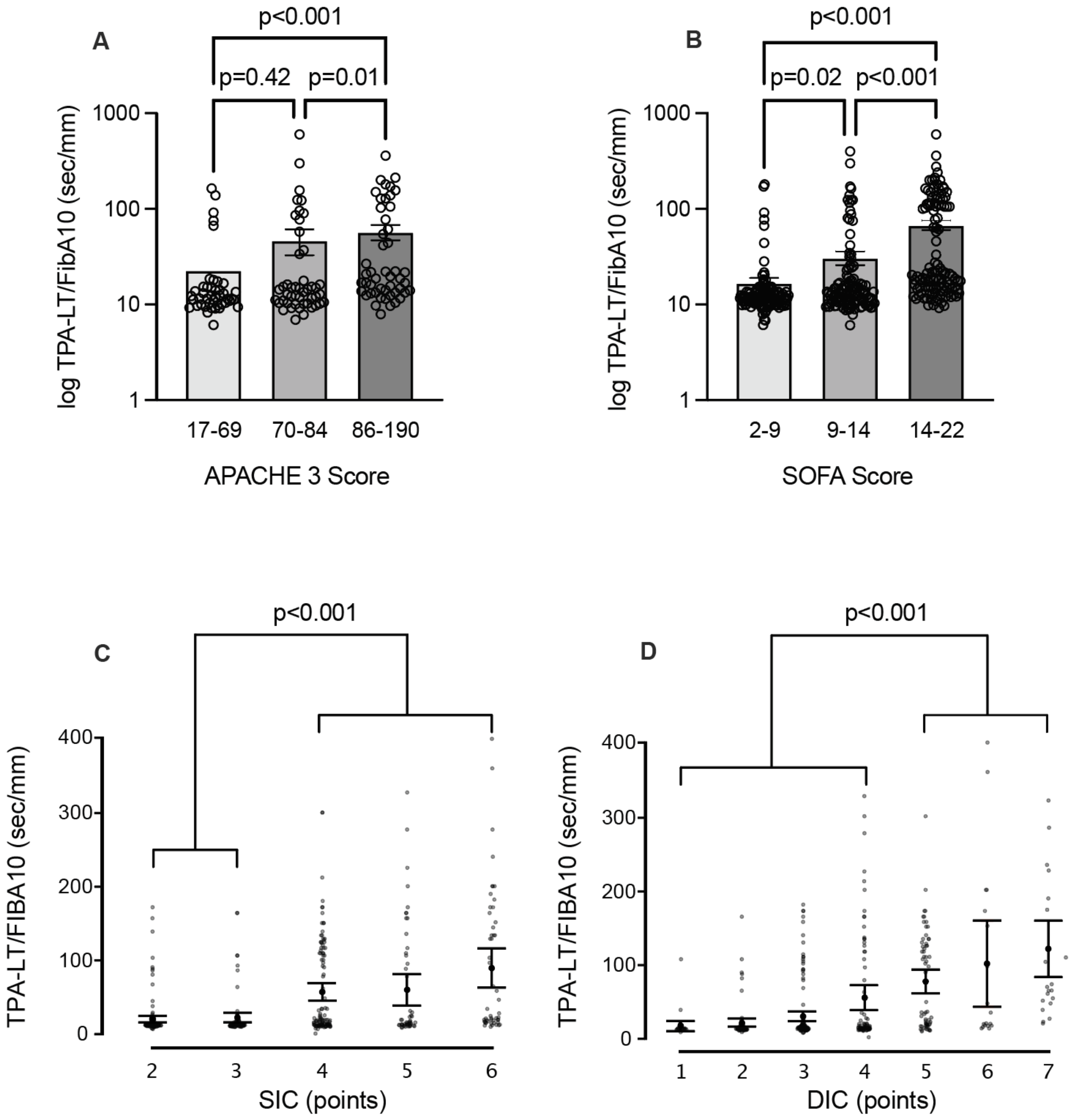

**Figure E5.** The predicted risk of death over the first week in ICU based on admission TPA-LT/FIBA10 ratio in the three grades of fibrinolysis resistance can be explored in the interactive data tool available at: https://fortitude.shinyapps.io/CHANGE_TPAFIB.

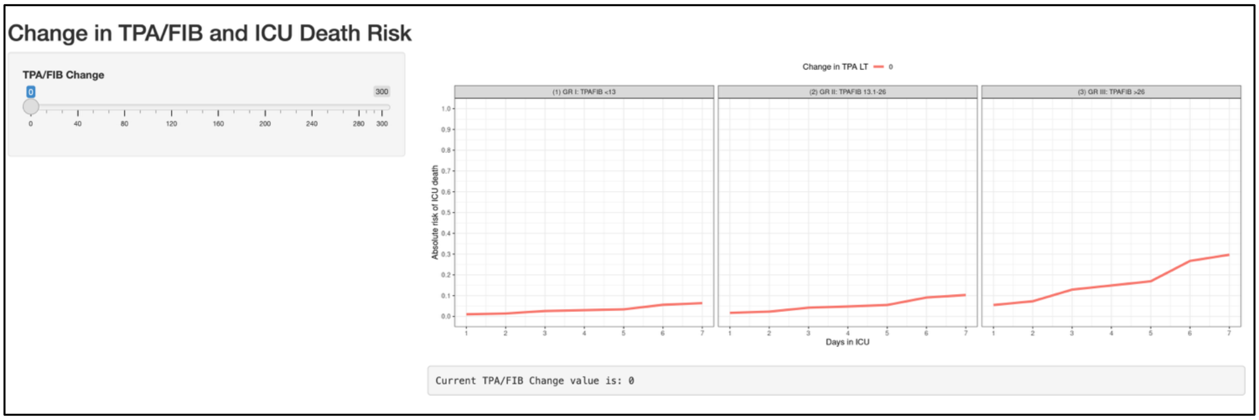


The slider is set to 0 sec/mm by default showing the impact of worsening fibrinolysis resistance as measured by increasing TPA-LT/FIBA10 by moving the slider to the right. Conversely, with the slider set to 300 sec/mm at the start, the impact of improving fibrinolysis resistance as measured by decreasing TPA-LT/FIBA10 is shown by moving the slider to the left.

**Detailed statistical methods.**

No formal sample size or power calculations were performed given the explorative nature of this study. The characteristics of study participants, ICU length of stay and ICU mortality, grouped by TPA-LT status at the time of admission to the ICU, are presented using descriptive statistics. Single missing variables to calculate the DIC score were replaced by multiple imputation used chained equations (MICE) with predictive mean matching [E1]. Standard laboratory coagulation variables and VET variables were used as predictors of missing values and convergence was assessed from trace plots showing stable estimates after 5 iterations. Results from 10 datasets were pooled and the score subsequently calculated. Correlations were analysed by the Spearman test. Cluster analysis was performed using a Gaussian Mixture Model. Values were offset to ensure all values were positive and a Box-Cox transformation was then applied with the optimal lambda parameter selected via maximum profile likelihood and normal distribution verified. The optimal number of clusters was set by the model based on the Bayesian Information Criterion. Cluster separation was characterised by the component means in the transformed space and cluster cutoffs were approximated by midpoints between sorted component means [E2]. Plasma fibrinolysis markers were analysed by a linear mixed-effects model using fibrinolysis resistance grade as a fixed effect and individual patients set as random effect. The degree of fibrinolysis resistance was compared across tertiles of APACHE III and SOFA scores, and across SIC and DIC scores with differences assessed using the Mann-Whitney U-test. Based on initial fibrinolysis resistance status, the probability of remaining in the ICU, leaving the ICU alive, or dying in the ICU up to 28 days was estimated using the Aalen-Johansen multistate approach [E3,E4]. The outcome classification was considered exhaustive, in that the combined estimates of the probabilities will sum to one for a given time interval [E3,E4]. A multi-state Markov model was used to estimate the probabilities of early transitioning through the fibrinolysis resistance states as well as into the absorbing states of death and discharge alive from ICU [E4,E5]. Patient demographics, initial TPA-LT/FIBA10 status, daily SOFA and APACHE III score were considered in the analysis as potential confounders and included in a Cox proportional hazards model [E6,E7]. The repeated TPA-LT/FIBA10 and SOFA scores were treated as time-varying covariates in the assessment of the independent association between change in TPA-LT/FIBA10 and risk of ICU mortality [E3,E4,E7]. The Cox proportional hazard coefficient for a change in TPA-LT/FIBA10 and the baseline hazards for ICU death were used to predict the risk of death for the first 7 days [E3,E4,E8]. The proportional hazards assumption in the Cox models was verified by visual inspection of smoothed Schoenfeld residual plots and a formal test of significance of an interaction term between time and TPA-LT/FIBA10 [E9]. The difference in the changes in TPA-LT/FIBA10 between ICU survivors and non-survivors was assessed using a random intercept and slope, linear mixed effect approach [E10]. Results are reported including the 95% confidence interval. In comparative analyses, the level of statistical significance was set at a two-sided p-value <0.05. Statistical analyses were performed using R 4.3.3 (R Core Team, 2024) including the 'mice', 'mclust', 'lmer', 'survival', 'survminer', 'nlme', 'cmprsk', 'msm', 'coxme' and 'rms' packages.

E3) Therneau TM, Grambsch PM. Modeling survival data : extending the Cox model. New York: Springer; 2000. xiii, 350 p.

E4) Kalbfleisch JD, Prentice RL. The statistical analysis of failure time data. 2nd ed. Hoboken, N.J.: J. Wiley; 2002. xiii, 439 p.

E5) Jackson C. Multi-State Models for Panel Data: The msm Package for R. Journal of Statistical Software. 2011;38(8):1-28.

E6) Cox DR. Regression models and life tables. J R Stat Soc (B). 1972;34:187-220.

E7) Therneau T, Crowson C, Atkinson E. Using Time Dependent Covariates and Time Dependent Coefficients in the Cox Model: Mayo Clinic Biostatistics Technical Report. 2024.

E8) Bradburn MJ, Clark TG, Love SB, Altman DG. Survival analysis part II: multivariate data analysis - an introduction to concepts and methods. Br J Cancer. 2003;89(3):431-6.

E9) Grambsch PM, Therneau TM. Proportional hazards tests and diagnostics based on weighted residuals. Biometrika 1994; 515-26.

E10) Pinheiro JC, Bates DM. Mixed effects models in S and S-PLUS. New York: Springer; 2000. xvi, 528 p.
